# ALLIANCE A022104/NRG-GI010: The Janus Rectal Cancer Trial: a randomized phase II/III trial testing the efficacy of triplet versus doublet chemotherapy regarding clinical complete response and disease-free survival in patients with locally advanced rectal cancer

**DOI:** 10.1101/2024.04.25.24306396

**Authors:** J Alvarez, Q Shi, A Dasari, J Garcia-Aguilar, H Sanoff, TJ George, TS Hong, G Yothers, PA Philip, GD Nelson, T Al Baghdadi, O Alese, W Zambare, DM Omer, FS Verheij, J Buckley, H Williams, M George, R Garcia, EM O’Reilly, JA Meyerhardt, A Shergill, N Horvat, PB Romesser, WA Hall, JJ Smith

## Abstract

**Background:** Recent data have demonstrated that in locally advanced rectal cancer (LARC), a total neoadjuvant therapy (TNT) approach improves compliance with chemotherapy and increases rates of tumor response compared to neoadjuvant chemoradiation (CRT) alone. They further indicate that the optimal sequencing of TNT involves consolidation (rather than induction) chemotherapy to optimize complete response rates. Data, largely from retrospective studies, have also shown that patients with clinical complete response (cCR) after neoadjuvant therapy may be managed safely with the watch and wait approach (WW) instead of preemptive total mesorectal resection (TME). However, the optimal consolidation chemotherapy regimen to achieve cCR has not been established, and a randomized clinical trial has not robustly evaluated cCR as a primary endpoint. Collaborating with a multidisciplinary oncology team and patient groups, we designed this NCI-sponsored study of chemotherapy intensification to address these issues and to drive up cCR rates, to provide opportunity for organ preservation, improve quality of life for patients and improve survival outcomes.

**Methods:** In this NCI-sponsored multi-group randomized, seamless phase II/III trial (1:1), up to 760 patients with LARC, T4N0, any T with node positive disease (any T, N+) or T3N0 requiring abdominoperineal resection or coloanal anastomosis and distal margin within 12 cm of anal verge will be enrolled. Stratification factors include tumor stage (T4 vs T1-3), nodal stage (N+ vs N0) and distance from anal verge (0-4; 4-8; 8-12 cm). Patients will be randomized to receive neoadjuvant long course chemoradiation (LCRT) followed by consolidation doublet (mFOLFOX6 or CAPOX) or triplet chemotherapy (mFOLFIRINOX) for 3-4 months. LCRT in both arms involves 4500 cGy in 25 fractions over 5 weeks + 900 cGy boost in 5 fractions with a fluoropyrimidine (capecitabine preferred). Patients will undergo assessment 8-12 (+/- 4) weeks post-TNT completion. The primary endpoint for the phase II portion will compare cCR between treatment arms. A total number of 296 evaluable patients (148 per arm) will provide statistical power of 90.5% to detect an 17% increase in cCR rate, at a one-sided alpha=0.048. The primary endpoint for the phase III portion will compare disease-free survival (DFS) between treatment arms. A total of 285 DFS events will provide 85% power to detect an effect size of hazard ratio 0.70 at a one-sided alpha of 0.025, requiring enrollment of 760 patients (380 per arm). Secondary objectives include time-to event outcomes (overall survival, organ preservation time and time to distant metastasis) and adverse effects. Biospecimens including archival tumor tissue, plasma and buffy coat in EDTA tubes, and serial rectal MRIs will be collected for exploratory correlative research. This study, activated in late 2022, is open across the NCTN and has a current accrual of 312. Support: U10CA180821, U10CA180882, U24 CA196171; https://acknowledgments.alliancefound.org.

**Discussion:** Building off of data from modern day rectal cancer trials and patient input from national advocacy groups, we have designed the current trial studying chemotherapy intensification via a consolidation chemotherapy approach with the intent to enhance cCR and DFS rates, increase organ preservation rates, and improve quality of life for patients with rectal cancer.

**Trial Registration:** Clinicaltrials.gov ID: NCT05610163; Support includes U10CA180868 (NRG) and U10CA180888 (SWOG)

## BACKGROUND

The use of total neoadjuvant therapy (TNT) is now at the forefront for patients with locally advanced rectal cancer (LARC) ^1–9^. The TNT treatment paradigm involves the delivery of both chemoradiation (CRT) and systemic chemotherapy in the neoadjuvant setting. There is mounting evidence that preoperative treatment leads to higher clinical and pathologic complete response (pCR) rates and improved treatment adherence, and TNT provides a unique opportunity to assess biological response on an individual patient basis. ^2,6,9,10^

As TNT has resulted in increased clinical complete response (cCR) rates, the need for surgery in patients with a cCR has been called into question, with increased interest in organ preservation (OP) and watch and wait (WW)/active surveillance strategies. It has long been known that patients with a pCR to preoperative CRT have lower tumor recurrence rates and improved survival compared to patients without a pCR, thus raising questions about the added value of total mesorectal excision (TME) for these individuals.^11–13^ Habr-Gama et al. were the first to report on the safety and efficacy of WW in patients with a cCR after CRT in 2004, noting 26% of patients were able to avoid surgery with durable complete response 10 years from chemotherapy and chemoradiation alone.^14^ Since then, multiple, large retrospective institutional case series and more recent prospective data suggest that WW can be safely incorporated without compromising oncologic outcomes. ^9,15^

The Organ Preservation in Patients with Rectal Adenocarcinoma (OPRA)^9^ trial was a prospective, multicenter phase II clinical trial in which patients with stage II/III rectal cancer were randomized to receive either induction long course chemoradiation (LCRT) followed by consolidative chemotherapy or induction chemotherapy followed by consolidative LCRT. Patients subsequently underwent TME or were offered surveillance via a WW protocol based on tumor response.^15^ The disease-free survival (DFS), overall survival (OS), local and distant recurrence-free survival were similar to patients treated with standard LCRT, TME, and adjuvant chemotherapy at both 3 and 5 years of follow-up.^9,16^ Approximately half of all patients treated with TNT achieved a cCR and were assigned to active surveillance rather than surgery. The use of induction LCRT followed by consolidation chemotherapy resulted in a higher rate of OP compared to induction chemotherapy followed by consolidative LCRT. ^9,16^

Here we report on the details of The Janus Rectal Cancer Trial (The Janus trial, NCT05610163), a National Clinical Trials Network (NCTN) trial testing the optimal TNT regimen using a consolidation chemotherapy approach of triplet versus doublet chemotherapy based on the hypothesis that a triplet chemotherapy regimen after induction LCRT will demonstrate superior cCR rates compared to a doublet chemotherapy regimen after induction LCRT. The Janus trial is important for our rectal cancer patients as it builds on the findings of modern rectal cancer trials to move the field forward in validation of the cCR endpoint and to enhance quality of life for patients through increased rates of organ preservation using a chemotherapy intensification TNT approach.^9,10^ During protocol development, the Janus study development team received input from two separate patient advocate groups and clinicians, noting that 76% of respondents preferred a chemotherapy intensification approach to a radiation escalation approach (Alvarez J, George M, Garcia R, et al. April 2024, *in review*). Based on OPRA data and patient input, we have designed the current trial studying chemotherapy intensification via a consolidation chemotherapy approach with the intent to enhance cCR and DFS rates, increase organ preservation rates, and thereby improve quality of life for patients with rectal cancer.

## METHODS

### Participants, interventions, and endpoints

#### Study Setting

The Janus Rectal Cancer Trial is organized through the Alliance for Clinical Trials in Oncology, sponsored by the National Cancer Institute (NCI) and administered through the NCTN. It is unique in that the study has integrated collaboration in both design and leadership across the NCI-NCTN inclusive of the Alliance for Clinical Trials in Oncology (overall PI and Chair, Smith), NRG Oncology (co-PI, Hall), SWOG (co-PI, Dasari), and ECOG (Study Champion, Alese). ClinicalTrials.gov Identifier: NCT05610163.

#### Patient selection and eligibility

Patients will be recruited and consented to the study in colorectal surgery, medical oncology, and radiation oncology clinics. In order to participate, patients must have a biopsy-proven clinical diagnosis of stage II or III (T4N0 or any T, node-positive disease) mismatch repair proficient adenocarcinoma of the rectum located 12 centimeters or less from the anal verge. Patients are only eligible if they have received no prior systemic chemotherapy, targeted therapy, immunotherapy, or radiation therapy administered as a treatment for colorectal cancer within the past five years and are older than 18 years old. Additional inclusion and exclusion criteria are provided in **Table 1**. Following informed consent, required eligibility testing will be completed, as well as a pelvic MRI with dedicated rectal protocol and a flexible sigmoidoscopy where the baseline tumor location and appearance will be documented, and a biopsy done. The patient’s eligibility checklist is verified by the local study team and then the patient is enrolled and randomized onto the trial.

**Table 1:**
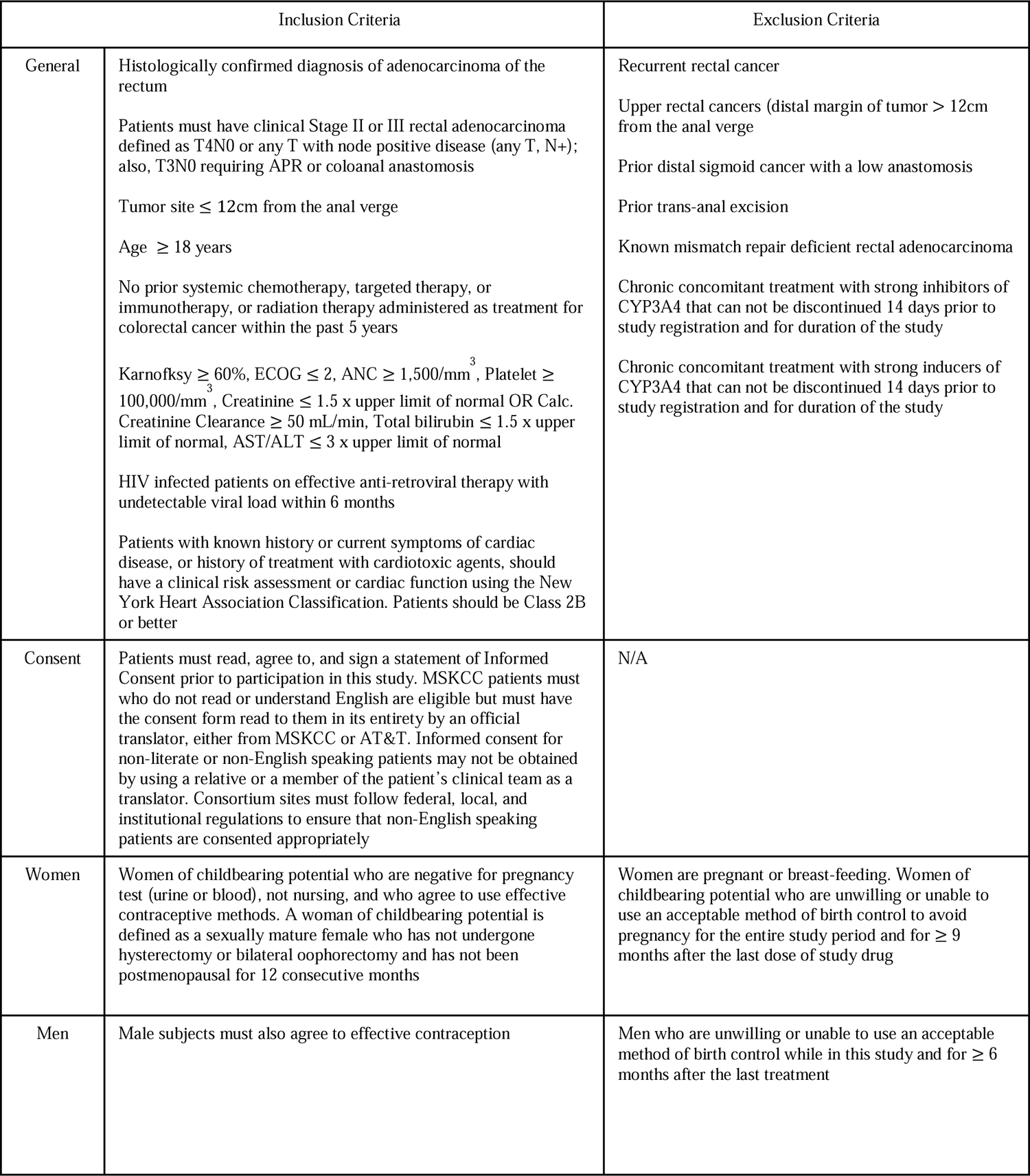
Inclusion and Exclusion Criteria.

| Inclusion Criteria |  | Exclusion Criteria |
| --- | --- | --- |
| General | Histologically confirmed diagnosis of adenocarcinoma of the rectum<br>Patients must have clinical Stage II or III rectal adenocarcinoma defined as T4N0 or any T with node positive disease (any T, N+); also, T3N0 requiring APR or coloanal anastomosis<br>Tumor site ≤ 12cm from the anal verge<br>Age ≥ 18 years<br>No prior systemic chemotherapy, targeted therapy, or immunotherapy, or radiation therapy administered as treatment for colorectal cancer within the past 5 years<br>Karnofsky ≥ 60%, ECOG ≤ 2, ANC ≥ 1,500/mm<sup>3</sup>, Platelet ≥ 100,000/mm<sup>3</sup>, Creatinine ≤ 1.5 x upper limit of normal OR Calc. Creatinine Clearance ≥ 50 mL/min, Total bilirubin ≤ 1.5 x upper limit of normal, AST/ALT ≤ 3 x upper limit of normal<br>HIV infected patients on effective anti-retroviral therapy with undetectable viral load within 6 months<br>Patients with known history or current symptoms of cardiac disease, or history of treatment with cardiotoxic agents, should have a clinical risk assessment or cardiac function using the New York Heart Association Classification. Patients should be Class 2B or better | Recurrent rectal cancer<br>Upper rectal cancers (distal margin of tumor > 12cm from the anal verge)<br>Prior distal sigmoid cancer with a low anastomosis<br>Prior trans-anal excision<br>Known mismatch repair deficient rectal adenocarcinoma<br>Chronic concomitant treatment with strong inhibitors of CYP3A4 that can not be discontinued 14 days prior to study registration and for duration of the study<br>Chronic concomitant treatment with strong inducers of CYP3A4 that can not be discontinued 14 days prior to study registration and for duration of the study |
| Consent | Patients must read, agree to, and sign a statement of Informed Consent prior to participation in this study. MSKCC patients must who do not read or understand English are eligible but must have the consent form read to them in its entirety by an official translator, either from MSKCC or AT&T. Informed consent for non-literate or non-English speaking patients may not be obtained by using a relative or a member of the patient's clinical team as a translator. Consortium sites must follow federal, local, and institutional regulations to ensure that non-English speaking patients are consented appropriately | N/A |
| Women | Women of childbearing potential who are negative for pregnancy test (urine or blood), not nursing, and who agree to use effective contraceptive methods. A woman of childbearing potential is defined as a sexually mature female who has not undergone hysterectomy or bilateral oophorectomy and has not been postmenopausal for 12 consecutive months | Women are pregnant or breast-feeding. Women of childbearing potential who are unwilling or unable to use an acceptable method of birth control to avoid pregnancy for the entire study period and for ≥ 9 months after the last dose of study drug |
| Men | Male subjects must also agree to effective contraception | Men who are unwilling or unable to use an acceptable method of birth control while in this study and for ≥ 6 months after the last treatment |

#### Study Design

The Janus Rectal Cancer Trial is a two arm, national, randomized, seamless phase II/III study investigating the effect of chemoradiation followed by either triplet chemotherapy or doublet chemotherapy in patients with LARC. The study was recently amended to power to create the definitive phase III DFS primary endpoint. The full study schema is illustrated in **Figure 1**.

**Figure 1:**
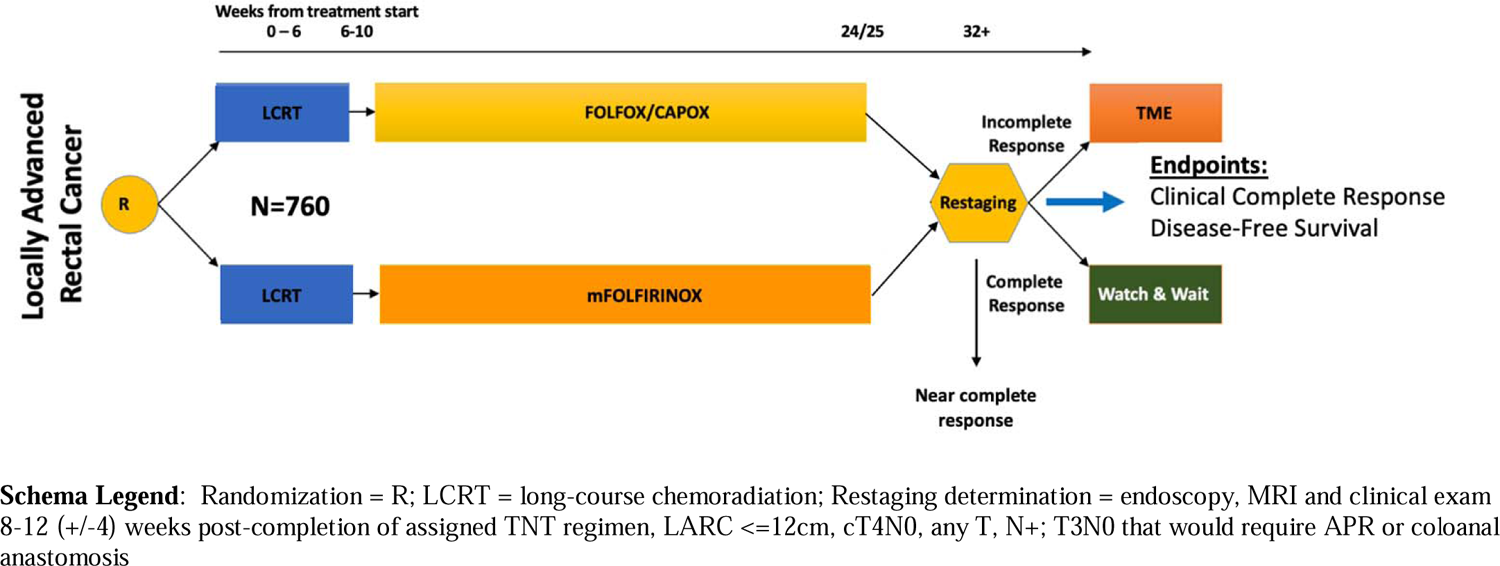
The Janus Rectal Cancer Trial Schema

#### Treatment Plan/Intervention

Protocol therapy will consist of induction LCRT followed by consolidation chemotherapy. Induction LCRT includes radiation (45Gy + 9 Gy boost in 27-30 fractions) in combination with a concomitantly administered fluoropyrimidine (preferred capecitabine; permissible substitution: continuous infusion 5-fluorouracil). Subsequently, patients will receive eight cycles of consolidative chemotherapy with either mFOLFOX6 (may be substituted by 5 cycles of CAPOX) in the control arm (Arm B) or eight cycles of mFOLFIRINOX in the experimental arm (Arm A). All patients will undergo assessment 8-12 (+/- 4) weeks post-completion of all therapy for the primary endpoint of cCR for the phase II portion. Patients who have an incomplete response will require TME, while patients who achieve a cCR will be recommended further management with WW. Uniquely, patients with a near complete response (nCR) will be recommended repeat assessment in 4-8 weeks and offered WW versus TME depending on their final response. If the tumor fails to evolve to a cCR then they will be recommended TME.

#### Primary Endpoints

The primary endpoints of the Janus Rectal Cancer Trial are to compare cCR rates and DFS between the two treatment groups for phase II and III portions, respectively. For the Phase II portion, the cCR rate is defined as the proportion of patients who achieved cCR at the end of TNT or who progressed to a cCR after nCR and re-evaluation. For the phase III portion, DFS is defined as time from date of randomization to the date of first occurrence of death due to all causes, tumor that recurs locally after an R0 resection TME, tumor that regrows after an initial apparent clinical and radiological complete response and cannot be surgically removed with an R0 resection TME, and/or M1 disease diagnosed at any point after the initiation of treatment. Note that local tumor regrowth that can be surgically removed with a R0 resection TME will not be a DFS event.

#### Secondary Endpoints

Secondary endpoints include organ-preservation-time, time to distant metastasis, OS, and rate of adverse events (AEs). Organ-preservation time is defined as time from the date of randomization to the date of the first occurrence of TME (including successful or attempted and failed TME), tumor that regrows after an initial apparent clinical and radiological complete response, and death due to all causes. Time to distant metastasis is defined as time from the date of randomization to the date of first documented distant metastasis. OS is defined as time from the date of randomization to the date of death due to all causes. The rate of AEs is defined as the proportion of patients experienced at least one Grade 3, Grade 4, or Grade 5 of each type of AE.

#### Exploratory Objective

Circulating tumor DNA (ctDNA) will be obtained from patients with consent during neoadjuvant therapy and surveillance with the aim to correlate values with radiographic, pathologic, and clinical outcomes.

The field of ctDNA assay development is rapidly evolving. Our study team will encourage prospective tissue and blood banking to then select the most appropriate assay based on sample availability and performance characteristics closer to the end of full study enrollment. Further details on biobanking protocol are included in the full protocol included in supplementary material.

#### Participant Timeline

Laboratory and clinical parameters during treatment are to be followed using individual institutional guidelines and the best clinical judgment of the responsible physician. It is expected that patients on this study will be cared for by physicians experienced in the treatment and supportive care of patients on this trial.

#### Pre-study Testing Intervals

The pre-study testing intervals are guidelines only. When calculating days of tests and measurements, the day a test or measurement is done is considered Day 0. Therefore, if a test were done on a Monday, the Monday one week later would be considered Day 7.

- To be completed ≤ 28 DAYS before registration: All laboratory studies, history and physical, performance status, pregnancy test.
- To be completed ≤ 42 DAYS before registration: Any X-ray, scan of any type or ultrasound which is utilized for tumor measurement per protocol.
- To be completed ≤ 60 DAYS before registration: Any baseline exams used for screening, or any X-ray, scan of any type or ultrasound of uninvolved organs which is not utilized for tumor measurement.

Please refer to **Table 2** for the complete Study Calendar for both arms in “Figures, tables and additional files.”

**Table 2:**
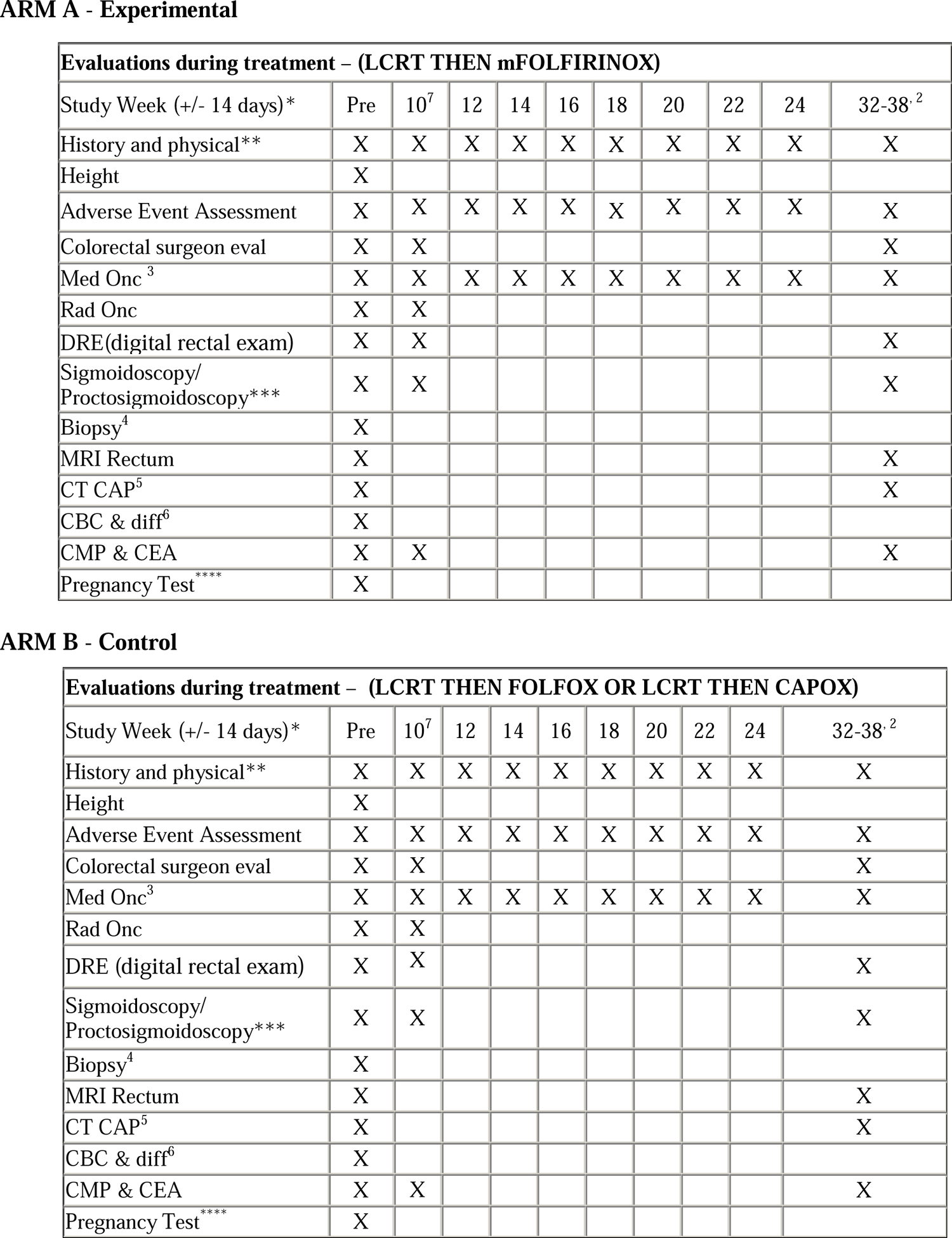
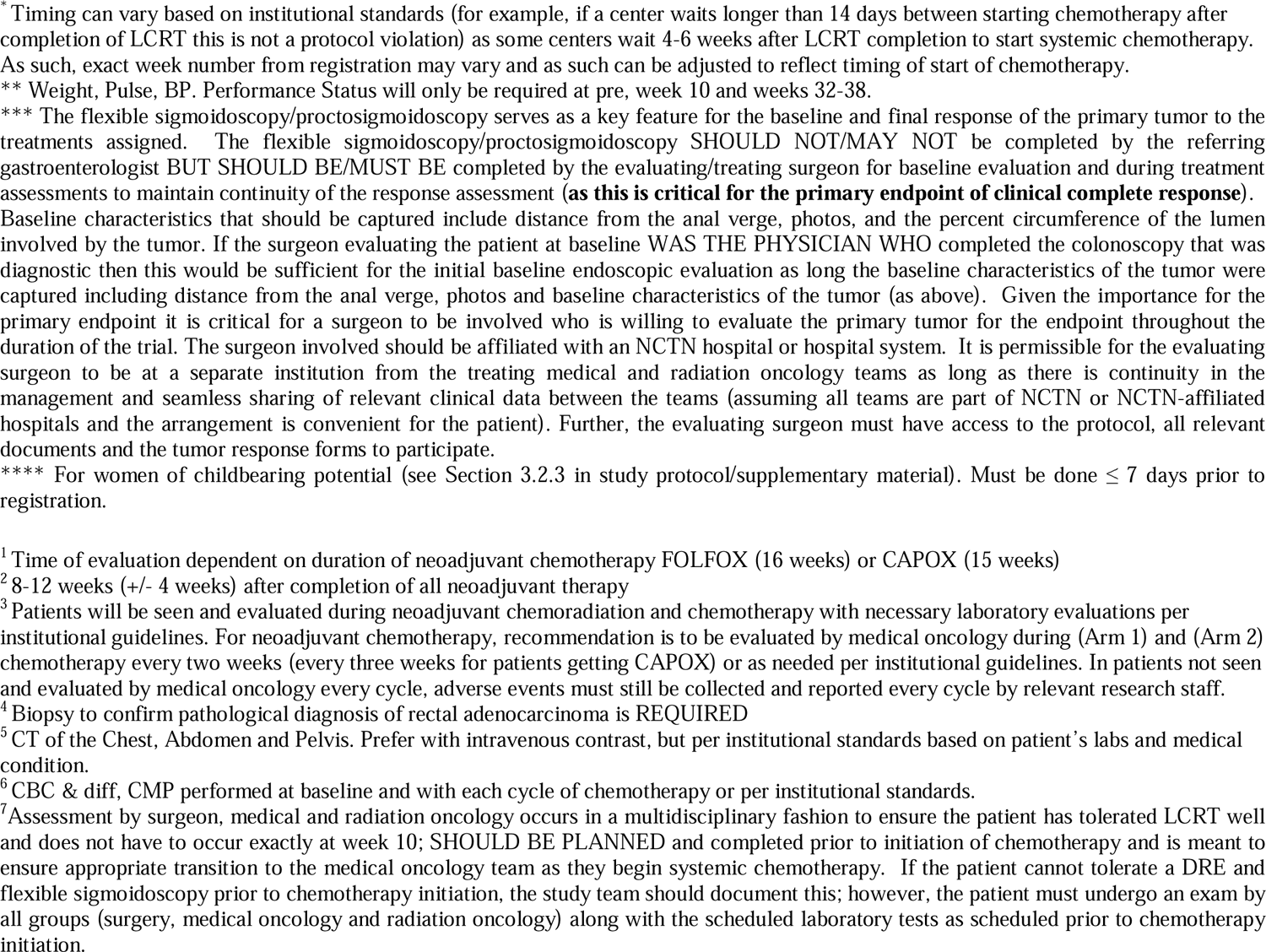
Study Calendars.

#### Sample Size

For the phase II portion, a total of 296 evaluable patients (148 per arm) will be needed to evaluate cCR rate. An additional 16 patients (5% inflation) will be accrued to account for cancelation after randomization and major violations. The total target accrual will be up to 312 patients.

For the phase III portion, Total sample size is 760 patients, or 380 per arm. Estimated accrual rate is 180 patients per year. Accrual as of April 19^th^ is 312.

### Assignment of Interventions

#### Randomization and stratification factors

Consenting and eligible patients will be registered to the study. Stratification factors will be recorded including (1) clinical tumor stage (T4 versus T1-3), (2) clinical nodal stage (N+ versus N0), and (3) distance from the lower edge of the tumor to the anal verge (0 to < 4cm; ≥ 4cm to < 8cm; ≥ 8cm to ≤12cm). Patients will be randomly assigned in a 1:1 ratio to one of the following treatment groups:

1. Induction LCRT followed by consolidative mFOLFIRINOX (Arm A) – experimental or study arm
2. Induction LCRT followed by consolidative mFOLFOX6 or CAPOX (Arm B) – control arm

#### Statistical Methods

The Phase II portion of this trial implements a group sequential design with a single interim analysis for futility evaluation, adopting Rho family (Rho=2) beta spending function for controlling the overall type II error rate.

The OPRA trial reported 52.4% (87 out of 166 randomized to consolidation chemotherapy arm) of patients who achieved a sustained cCR and preserved the rectum. For the proposed trial, we assume a cCR rate of 50% in the control arm (Arm B). A total number of 296 evaluable patients will provide 90.5% power to detect an 17% increase in cCR rate (67% in the experimental arm [Arm A]) at a one-sided type I error rate of 0.048.

The Phase III portion implements a group sequential design with one futility interim analysis based on a non-binding beta spending function (Rho family with Rho=3.2), which will be performed when 50% of DFS events have been observed (143 events). The OPRA trial reported a three-year DFS rate of 76% (95% CI, 69-83%) for patients who received CRT followed by consolidation chemotherapy.^9^ A total number of 285 DFS events will provide 85% power to detect an effect size of hazard ratio (HR) = 0.70 (3-year DFS rate of 82.5% in the experimental arm A) at a one-sided type I error rate of 0.025. With further assumptions of an accrual rate of 180 patients per year and a minimum of four years of follow-up, a maximum of 760 patients (380 in each arm) are required to enroll, unless the study team makes a decision of early termination (monitoring rules specified in supplementary protocol).

For the phase II primary endpoint of cCR rate, hypothesis testing will be performed on the modified intent to treat population defined as all patients properly randomized, completed LCRT and who started at least one dose of protocol defined chemotherapy treatment, with treatment grouping according to the original assignment at randomization. Sensitivity analysis will be formed on the per protocol population defined as all patients properly randomized who started at least three cycles of chemotherapy after LCRT, with treatment grouping according to actual treatment received during the first cycle of chemotherapy. An interim analysis for futility will be performed when 50% of patients in each arm (74 patients) are randomized and cCR status is determined. The analysis of the phase III primary endpoint of DFS will be performed on Intention-to-treat population (ITT) defined as all patients who are properly randomized, regardless of the actual treatment received. The treatment grouping will be according to the original assignment at randomization. Sensitivity analyses will be performed on mITT and PP population. At interim and final analyses, stratified Cox model will be conducted to compare DFS in the experimental arm to DFS in the control arm with stratification factors as stratum, based on all data collected at the analysis time point.

The analysis of secondary endpoints will be on mITT and PP population with the Kaplan-Meier method and stratified Cox regression models. The maximum grade for each type of AE related to study treatment will be recorded and reviewed to determine patterns. The overall AE rates for grade 3 or higher AEs will be compared between two treatment groups using Chi-square test (or Fisher’s exact test if the data in the contingency table is sparse).

### Monitoring

#### Response evaluation

Patients will undergo assessment for tumor response at 8-12 (+/- 4) weeks post-completion of TNT. Patients with a cCR as determined by the MSK regression schema^9,15^ (no tumor on clinical exam, endoscopy, or MRI) may be offered a WW approach or TME depending on the outcome of an in-depth discussion and understanding of the risks and benefits of each approach. Patients with an incomplete response as determined by the MSK regression schema (any evidence of residual tumor on clinical exam, endoscopy, or MRI) will be recommended to undergo a TME. Similar to guidelines in the OPRA Trial, if patients have a near complete response (nCR) they can undergo repeat assessment 4-8 weeks later. If there is evidence the tumor has stopped responding, continues to persist, or regrows then the patient will be recommended to undergo a TME. Endoscopy will be the deciding factor on determination of cCR if there is a discrepancy between clinical exam and MRI findings.

#### Neoadjuvant treatment completion monitoring

The completion of neoadjuvant treatment will be closely monitored. We will compare early off treatment (EOT) rates between both treatment arms at select timepoints. If the difference in EOT rate (experimental arm minus control arm) is greater than specified thresholds, a formal review will be triggered and potential protocol modifications, including possible halting of accrual, will be formulated under the consultation with Cancer Therapy Evaluation Program (CTEP).

#### R0 resection for patients on WW/active surveillance monitoring

We will carefully monitor the R0 resection rate among patients who proceed to a WW strategy after TNT and later require TME during follow-up. Patients enrolled on both arms will be pooled for this monitoring. We have employed specific monitoring rules to test the hypothesis of whether an R0 resection rate in our population is adequate.

#### Tumor Regrowth

We will closely monitor tumor regrowth in patients who proceed to WW strategy after TNT. The five-year follow up data from OPRA reported 29% regrowth rate in patients randomized to induction LCRT and consolidation chemotherapy group who proceeded to a WW strategy.^16^ The one-year regrowth rate will be defined as number of patients who experience tumor regrowth (regardless of whether it can be salvaged by TME) within one year after the last dose of pre-operative TNT divided by the total number of patients in the analysis population. The one-year regrowth rate will be estimated within each arm separately, when all patients in this population are followed for at least one year after last dose of chemotherapy.

#### Patient safety monitoring

The Study Chair(s) and the Study Statistician will review the study monthly to identify accrual, AE/safety trends, and any emerging concerns. The Study team will have monthly meetings to identify any issues that arise during the Phase II and Phase III portions of the study.

### Disease Evaluation

#### Measurement of Treatment Effect

Follow up after treatment consists of a schedule of endoscopy, digital rectal exam (DRE), CT Chest/Abdomen/Pelvis, MRI Pelvis, and CEA (**Table 3**). For specific surveillance intervals, refer to **Table 4a and 4b** for patients following WW protocol or for patients who are post-TME.

**Table 3:** Protocol Duration of Follow Up.

|  | **MRI rectum** | **Endoscopy and digital rectal exam** | **CT chest, abdomen, pelvis** | **CEA** |
| --- | --- | --- | --- | --- |
| All patients who completed pre-operative protocol treatment | Pre-treatment, post-TNT re-staging | Pre-treatment, post-TNT re-staging | Pre-treatment, post-TNT re-staging, annually from 5 years post-registration | Pre-treatment, every 3-6 months post-TNT in Years 1-2, every 6 months post-TNT in Years 3-5 |
| All patients who are off pre-operative protocol treatment early due to Progression of disease- | As per treatment team | As per treatment team | As per treatment team | As per treatment team |
| All patients who are off pre-operative protocol treatment early due to other reasons (patient refusal, clinician decision to withdraw/other) | As per treatment team | As per treatment team | As per treatment team | As per treatment team |
| Patients in WW group | After post-TNT re-staging: Q6 months in Years 1-2, Q12 months in Years 3-5 (As clinically indicated years 4-5) | Post-TNT re-staging: Q3 months in Years 1-2, Q6 months in Years 3-5 | Pre-treatment, post-TNT re-staging, annually from 5 years post-registration | Pre-treatment, every 3-6 months post-TNT in Years 1-2, every 6 months post-TNT in Years 3-5 |

**Table 4:** Evaluations during surveillance for WW and post-TME.

| Table 4a: Evaluations during follow-up for WW patients (after completion of TNT with cCR and post-TNT restaging) |  |  |  |  |  |  |  |  |  |  |  |  |  |  |
| --- | --- | --- | --- | --- | --- | --- | --- | --- | --- | --- | --- | --- | --- | --- |
| Years on study | Year 1<br>(Every 3 months) |  |  |  | Year 2<br>(Every 3 months) |  |  |  | Year 3<br>(Every 6 months) |  | Years 4-5**<br>(Every 6 months) |  |  |  |
| Months after post-TNT restaging (+/- 30 days) | 3 | 6 | 9 | 12 | 15 | 18 | 21 | 24 | 30 | 36 | 42 | 48 | 54 | 60 |
| History and Physical | X | X | X | X | X | X | X | X | X | X | X | X | X | X |
| Sigmoidoscopy/<br>Proctosigmoidoscopy | X | X | X | X | X | X | X | X | X | X | X | X | X | X |
| MRI Rectum |  | X |  | X |  | X |  | X |  | X |  | X |  | X |
| CT CAP<sup>1</sup> |  |  |  | X |  |  |  | X |  | X |  | X |  | X |
| CEA levels<sup>2</sup> | X | X |  | X | X | X |  | X | X | X | X | X | X | X |

\*If TME completed via abdominoperineal resection – not relevant and unnecessary
\*\*MRI in years 4 through 5 would only be supported if clinically indicated by treating team.
<sup>1</sup> CT of the Chest, Abdomen and Pelvis if regrowth occurs and TME completed then patients will be followed as per NCCN guidelines
<sup>2</sup> After 24 months, CEA will be evaluated every 6 months up to five years, based on NCCN guidelines.

| Table 4b: Evaluations during follow-up for TME patients (after definitive surgical resection)<sup>#</sup> |  |  |  |  |  |  |  |  |  |  |  |  |  |  |
| --- | --- | --- | --- | --- | --- | --- | --- | --- | --- | --- | --- | --- | --- | --- |
| Years on study | Year 1<br>(Every 3 months) |  |  |  | Year 2<br>(Every 3 months) |  |  |  | Year 3<br>(Every 6 months) |  |  | Years 4-5**<br>(Every 6 months) |  |  |
| Months after treatment (+/- 30 days) | 3 | 6 | 9 | 12 | 15 | 18 | 21 | 24 | 30 | 36 | 42 | 48 | 54 | 60 |
| History and Physical | X | X | X | X | X | X | X | X | X | X | X | X | X | X |
| Sigmoidoscopy/<br>Proctosigmoidoscopy |  |  |  | X |  |  |  | X |  | X |  | X |  | X |
| CT CAP<sup>1</sup> |  | X |  | X |  | X |  | X |  | X |  | X |  | X |
| CEA levels<sup>2</sup> | X | X | X | X | X | X | X | X | X | X | X | X | X | X |

<sup>#</sup>Institutional guidelines for surveillance can be followed post-TME in accordance with NCCN guidelines or as clinically indicated per the discretion of the treating physicians
\*If TME completed via abdominoperineal resection – A FLEXIBLE SIGMOIDOSCOPY/PROCTOSIGMOIDOSCOPY IS not relevant and unnecessary to assess the anastomosis for recurrence
<sup>1</sup> CT of the Chest, Abdomen and Pelvis: TME group patients will be followed according to NCCN guidelines.
<sup>2</sup> After 24 months, CEA will be evaluated every 6 months up to five years, based on NCCN guidelines.

#### Clinical Tumor Evaluation

On endoscopy, the length of the tumor is defined as the difference between the distance of the proximal and distal margins in relation to the anal verge. Endoscopic tumor response will be determined by the MSK Regression Schema (**Table 5**). For patients who ultimately undergo TME after TNT, clinical tumor evaluations with DRE, endoscopy, and MRI will determine the need for TME. For patients who elect for a WW approach, clinical tumor evaluations with DRE, endoscopy, MRI, CT Chest/Abdomen/Pelvis, and CEA will occur during the post-TNT follow-up, up to 5 years post randomization, or up to salvage TME, whichever occurs first (**Table 3**).

**Table 5:** The MSK Regression Schema, Please refer to the following video for endoscopic and MRI response assessment - https://www.youtube.com/watch?v=38rsqZvJIHg.

| **MSK Regression Schema<sup>#</sup>** |  |  |  |
| --- | --- | --- | --- |
|  | **Clinical Complete Response** | **Near Complete Response** | **Incomplete / No Response** |
| **Endoscopy** | Flat, white scar<br>Telangiectasia<br>No ulcer<br>No nodularity | Irregular mucosa<br>Superficial ulceration<br>Mild persisting erythema of the scar | Visible tumor |
| **Digital Rectal Exam** | Normal<sup>^</sup> | Smooth induration or minor mucosal irregularity<sup>^</sup> | Palpable tumor nodules<sup>^</sup> |
| **MRI-T2WI** | Normal appearing rectal wall<br>OR<br>Only fibrosis (dark T2 signal) and no intermediate signal intensity at the site of tumor<sup>*</sup><br>AND<br>No suspicious lymph nodes<sup>**</sup> | Predominantly fibrosis at the site of tumor<sup>*</sup> with punctate areas of T2 intermediate signal<br>AND/OR<br>No suspicious or borderline enlarged lymph nodes<sup>**</sup> | Predominantly residual tumor with T2 intermediate signal and no or minimal fibrosis at the site of tumor<sup>*</sup><br>AND/OR<br>Suspicious lymph nodes<sup>**</sup><br>AND/OR<br>Mucin at the site of tumor<sup>****</sup> |
| **MRI-DWI** | No restricted diffusion<sup>***</sup> at the site of tumor<sup>*</sup> | Punctate areas of restricted diffusion<sup>***</sup> at the site of tumor<sup>*</sup> | Restricted diffusion<sup>***</sup> at the site of tumor<sup>*</sup> |

\* Site of tumor: rectal wall, extramural vascular invasion and/or tumor deposit.
\*\* Suspicious lymph nodes criteria: (a) mesorectal and superior rectal nodes > 0.5 cm in the short axis; (b) internal iliac > 0.4 cm in the short axis particularly if suspicious on baseline; (c) obturator > 0.6 cm in the short axis particularly if suspicious on baseline; (c) mucin within the lymph nodes since MRI cannot distinguish cellular from acellular mucin. Additional lymph nodes should be interpreted cautiously, as there are no well-defined radiological criteria to strongly support their significance.
\*\*\* Restricted diffusion: high signal on DWI high b-value (minimum b800) and low signal on ADC map.
*T2 dark through* (low signal on both DWI and ADC map) and *T2 shine through* (high signal on both DWI and ADC map) effects are not considered restricted diffusion.
\*\*\*\* MRI is unable to differentiate cellular from acellular mucin.

#### The MSK Regression Schema

The MSK Regression Schema (**Table 5**) is based on subjective endoscopic and radiologic findings.^9,15^ It was developed by consensus with the aid of expert colorectal surgeons, medical oncologists, radiation oncologists, radiologists, and pathologists prior to the start of the OPRA trial to serve as a guideline to assess response and to provide uniformity in determining cCR, nCR, and incomplete/no tumor response after a patient has completed TNT (**Figure 2 and 3**).

**Figure 2:**
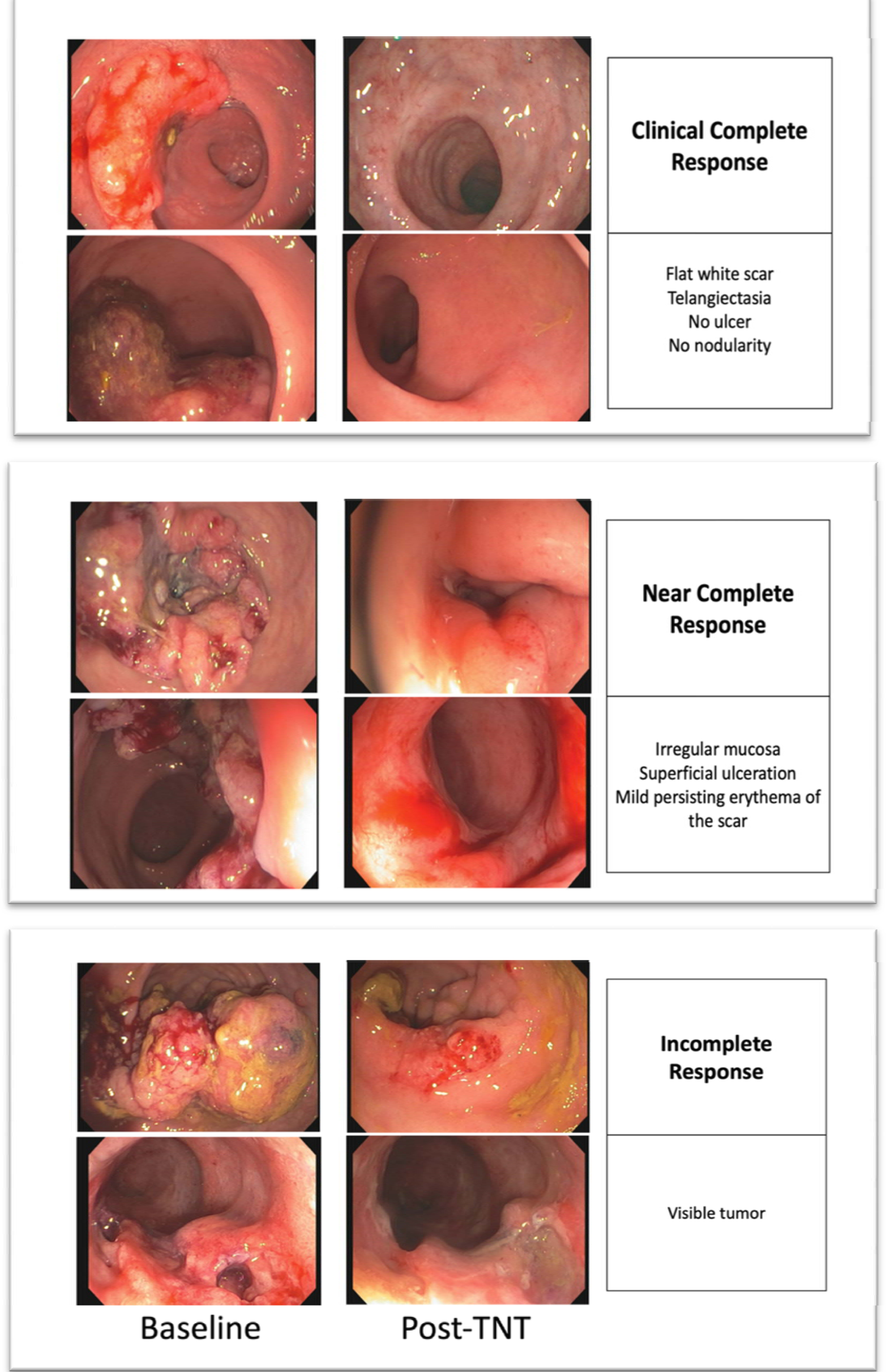
Endoscopy images showing a clinical complete, near complete, and incomplete response for patients who have completed TNT.

**Figure 3:**
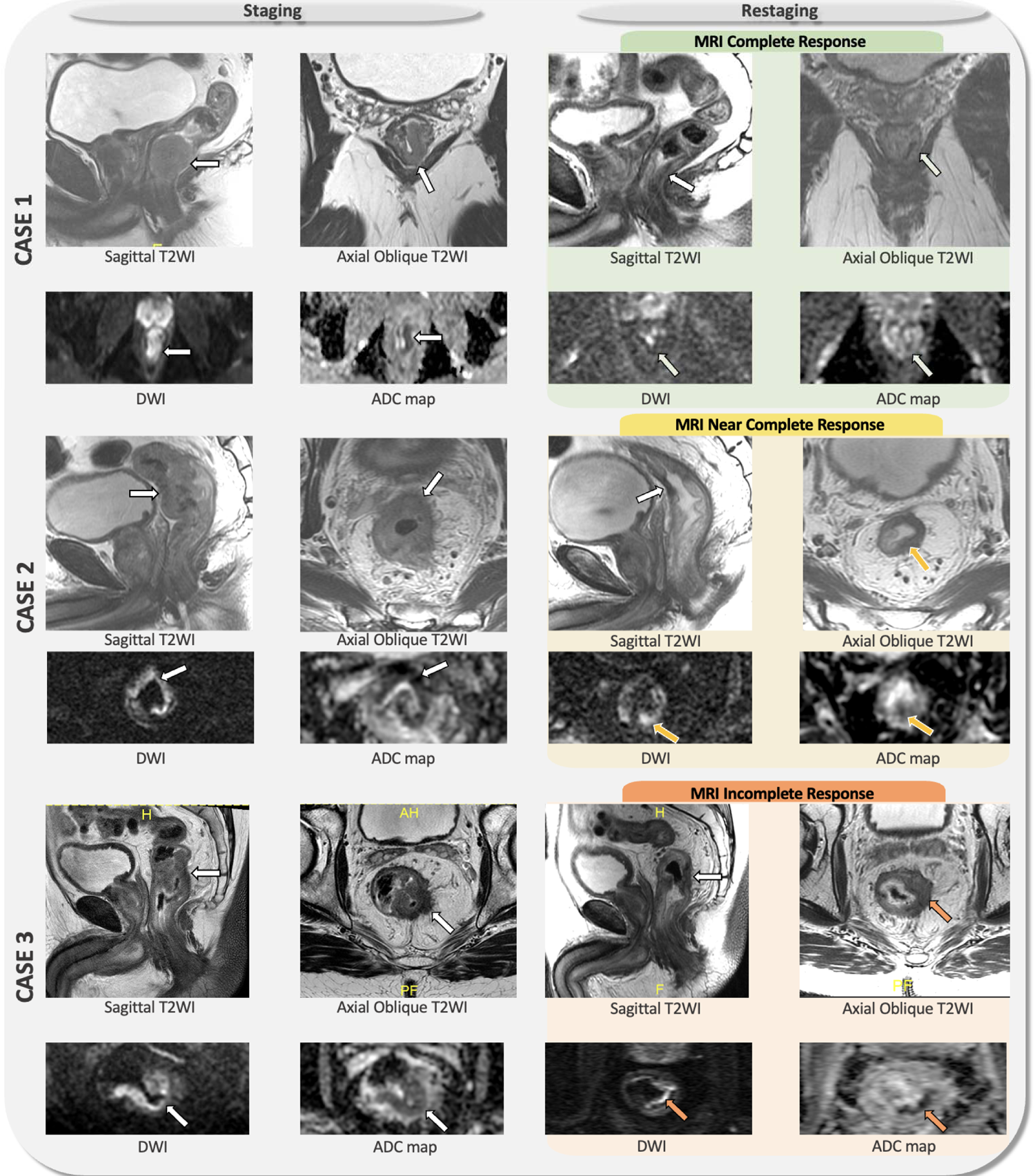
MRI images showing baseline and restaging MRI rectum of patients with complete response (case 1), near complete response (case 2) and incomplete response (case 3) based on the MRI assessment. Reviewing the baseline MRI is highly recommended, since it is helpful to guide how to provide the best oblique T2 angulation and to locate the tumor bed. T2-weighted imaging (T2WI) and diffusion-weighted imaging (DWI) are the sequences used for imaging interpretation. Clinical complete response on MRI (green) is characterized by normal appearing rectal wall or only fibrosis (low signal intensity on T2WI) an no areas of viable tumor (intermediate signal intensity on T2WI) without areas of restricted diffusion in the tumor bed. Near complete response on MRI (yellow) is defined as very small area of viable tumor with punctate restriction on DWI (high signal intensity on DWI and low signal intensity on ADC map) or borderline lymph nodes. Finally, incomplete response on MRI (orange) represents significant areas of residual tumor.

#### Radiologic Tumor Evaluation

Standard T2-weighted imaging (T2WI) and diffusion-weighted imaging (DWI) sequences will be obtained in 1.5T or 3.0T units using phased-array body coil. Expert radiologists from the patient’s primary treatment center will interpret all imaging studies according to the MSK regression schema.^15^ Patients will require a baseline MRI and re-staging evaluations with MRI will be required of patients within the WW group (every 6 months for years 1-2, and every 12 months for years 3-5). Central radiology review is not required, but imaging data will be centrally collected. Refer to **Table 6** which describes MRI features associated with local regrowth.

**Table 6:**
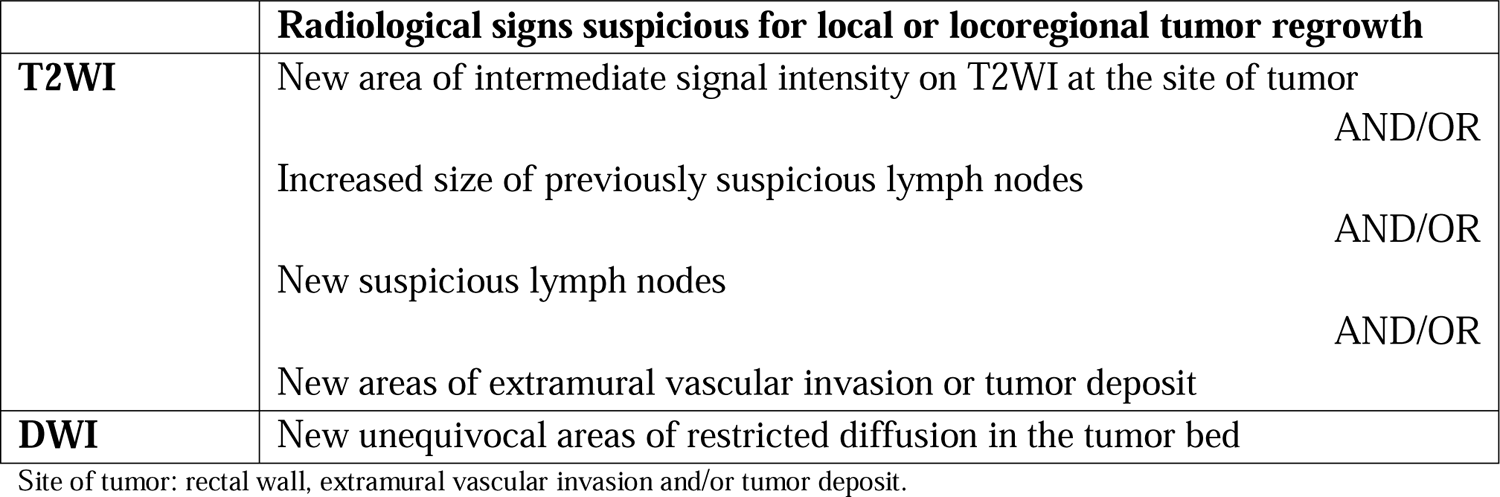
Radiologic Features suspicious for Local or locoregional tumor Regrowth on MRI.

#### Protocol Follow Up

Protocol intervention will continue until completion of LCRT and consolidation chemotherapy (8 cycles of FOLFOX or 5 cycles of CAPEOX), local/distant disease progression which preclude surgery, or unacceptable AEs. Patients who proceed to a WW strategy will be monitored as described in **Table 3** for up to five years. TME will be performed as appropriate.

## DISCUSSION

The Janus Rectal Cancer Trial expands on the findings of modern rectal cancer trials ^4,9,10,15^ to provide further evidence for cCR as an endpoint and demonstrate improved patient outcomes with a consolidation chemotherapy intensification TNT approach. Preserving the rectum is a significant quality of life benefit for those patients who achieve a cCR and can progress to WW/active surveillance as it spares patients the morbidity of radical surgery and potential long term sequelae.^17–20^ In addition, this trial will allow a venue to prospectively validate the schema used in OPRA for assessing tumor response, and allow us to gain critical insight into the biology of response to consolidation TNT approaches in the context of standard clinical measures and novel correlative biomarkers.

Patients in the Janus trial are randomized to induction LCRT followed by either mFOLFOX6/CAPEOX (doublet chemotherapy) or mFOLFIRINOX (triplet chemotherapy). Patients either proceed to surgery or WW based on tumor response. Multiple phase II and phase III clinical trials in metastatic colorectal cancer patients have compared doublet chemotherapy to triplet chemotherapy and have found consistently improved outcomes including objective radiographic response rates, OS and profession-free survival (PFS). ^21–28^ Based on these results, the triplet regimen is included among first-line options in most clinical guidelines and recommendations worldwide. ^29–31^ More recently, the PRODIGE-23 trial enrolled 460 patients with LARC and randomized them to pre-operative CRT, TME, and adjuvant FOLFOX (control arm) versus induction mFOLFIRINOX followed by CRT, TME, and adjuvant FOLFOX (experimental arm).^4,10^ The addition of 6 cycles of neoadjuvant mFOLIRINOX prior to CRT increased the pCR from 12 to 27%. Importantly, the 7-year updated data from PRODIGE-23 presented at ASCO 2023 demonstrated significantly better DFS, metastasis-free survival, and OS in the triplet TNT arm versus control arm (68% vs. 63% DFS). Together, these data convincingly show increased efficacy of triplet over doublet chemotherapy in patients with advanced colorectal cancer in improving R0 resection rates, objective response rates, PFS, DFS, and OS

The Janus trial will expand on the findings from OPRA demonstrating improved organ preservation rates utilizing a consolidation chemotherapy approach.^9^ The 5-year updated data in the OPRA trial has since resulted and demonstrated stable organ preservation rates for the consolidation chemotherapy arm (54%) versus the induction chemotherapy arm (39%, p=0.012). Further, local regrowth rates remained lower in the consolidation chemotherapy arm (29%) versus in the induction chemotherapy arm (44%) (p=0.02).^16^ The TIMING trial^7^ reported increased pCR rates with additional cycles of FOLFOX in the consolidation setting. Additional evidence for the efficacy of the consolidation chemotherapy approach has been shown in the recent German trial/ CAO/ARO/AIO-12 with acceptable pCR and superior complete response rates (25% pCR vs 17% pCR) compared to an induction chemotherapy approach ^6,32^. Building on these data and the data from OPRA trial, we anticipate that employing FOLFIRINOX in a consolidation approach after LCRT has the potential to further drive response rates higher (increasing cCR rates) with an associated increase in organ preservation rates.

Lastly, this study will measure ctDNA levels and study potential use as an exploratory biomarker in the context of a prospective randomized trial. ctDNA levels will be used to measure response to treatment and may become a useful tool to help patients and clinicians choose TME versus WW. We also aim to develop a minimal residual disease-based risk classification for cCR patients. ctDNA has shown promise especially in the realm of colorectal cancer.^33–38^ Multiple groups have reported worse recurrence-free survival in patients with positive ctDNA post CRT, further supporting it’s utility during surveillance.^33–36, 39^ Despite promising preliminary data, the kinetics of ctDNA after TNT, TME, and during surveillance and its correlation with disease recurrence and overall survival has not been adequately studied. Our study serves as the optimal platform to study ctDNA as a predictive and prognostic biomarker in locally advanced rectal cancer.

A major criticism of previously published WW data is that most of it comes from a select patient population treated at specialized centers. The recently reported OPRA trial was conducted across sixteen highly specialized academic centers, and thus provides the most robust, prospective data on outcomes of WW. While all OPRA sites were selected based on the expertise and clinical interest of the surgical team, the Janus trial will determine generalizability of a WW approach across a more diverse population of patients, practice sites, and providers while incorporating a chemotherapy intensification approach in the context of modern TNT to improve response outcomes in a seamless phase II/III trial incorporating the WW strategy in a manner that will be acceptable to patients and clinicians. By running this trial through the NCI’s National Cancer Trial Network, Janus expands the opportunity to consider WW for patients treated at academic and community practices across the United States.

In summary, this study will explore the advantages of triplet versus doublet chemotherapy in LARC patients while expanding on findings from prior landmark trials by offering WW as an alternative to surgery in a national trial completed in a heterogeneous group of centers. We aim to optimize cCR rates via a chemo-intensification method, which was preferred by patients when surveyed in two separate patient advocate groups. In addition to cCR for the phase II portion, DFS will be the primary endpoint for our phase III portion. We will also evaluate and compare organ-preservation time, time to distant metastasis, OS, and toxicity profiles of TNT. Finally, we will conduct ctDNA surveillance and correlate it with patient outcomes, radiologic, and pathologic findings. Regardless of the trial results, this study has the potential to significantly impact the care of patients with locally advanced rectal cancer across the United States.

## DECLARATIONS

### Ethics approval and consent to participate

As of March 1, 2019, all U.S.-based sites must be members of the NCI Central Institutional Review Board (NCI CIRB) in order to participate in CTEP and Division of Cancer Prevention studies open to the NCTN and NCI Community Oncology Research Program (NCORP) Research Bases. In addition, U.S.-based sites must accept the NCI CIRB review to activate new studies at the site after March 1, 2019. Local IRB review will continue to be accepted for studies that are not reviewed by the CIRB, or if the study was previously open at the site under the local IRB. International sites should continue to submit Research Ethics Board (REB) approval to the CTSU Regulatory Office following country-specific regulations.

Refer to the protocol in supplementary material for additional information. The patient must be aware of the neoplastic nature of his/her disease and willingly consent after being informed of the procedure to be followed, the experimental nature of the therapy, alternatives, potential benefits, side-effects, risks, and discomforts. Current human protection committee approval of this protocol and a consent form is required prior to patient consent and registration. Patients with an impaired decision-making capacity may be enrolled in this study, where institutional policy and IRB of record allow.

### Consent for Publication

Appropriate consent was obtained for images used in this manuscript.

### Availability of data and materials

Not applicable

## Authors’ contributions

Authors with asterisk contributed equally to drafting the final version of the submitted manuscript. Each author contributed equally in revising the final version of the submitted manuscript.

## Data Availability

Not applicable, All data produced in the present study are available upon reasonable request to the authors

## Acknowledgements

https://acknowledgments.alliancefound.org

## LIST OF ABBREVIATIONS

Abbreviations: Definition

cCR: Clinical complete response

CRT: Chemoradiation

ctDNA: Circulating tumor DNA

CTEP: Cancer Therapy Evaluation Program

DFS: Disease-free survival

DRE: Digital rectal examination

LARC: Locally advanced rectal cancer

EOT: Early off treatment

LCRT: Long course chemoradiation

NCI: National Cancer Institute

nCR: Near complete response

NCTN: National Clinical Trials Network

OPRA: Organ Preservation in Patients with Rectal Adenocarcinoma

OS: Overall survival

PFS: Progression-free survival

pCR: Pathologic complete response

TME: Total mesorectal excision

TNT: Total neoadjuvant therapy

WW: Watch and wait approach/Active surveillance

## REFERENCES

1. Bahadoer RR, Dijkstra EA, van Etten B, et al. Short-course radiotherapy followed by chemotherapy before total mesorectal excision (TME) versus preoperative chemoradiotherapy, TME, and optional adjuvant chemotherapy in locally advanced rectal cancer (RAPIDO): a randomised, open-label, phase 3 trial. Lancet Oncol. 2021;22(1):29–42. doi:10.1016/S1470-2045(20)30555-6

2. Cercek A, Roxburgh CSD, Strombom P, et al. Adoption of Total Neoadjuvant Therapy for Locally Advanced Rectal Cancer. JAMA Oncol. 2018;4(6):e180071. doi:10.1001/jamaoncol.2018.0071

3. Chau I, Brown G, Cunningham D, et al. Neoadjuvant Capecitabine and Oxaliplatin Followed by Synchronous Chemoradiation and Total Mesorectal Excision in Magnetic Resonance Imaging– Defined Poor-Risk Rectal Cancer. J Clin Oncol. Published online September 21, 2016. doi:10.1200/JCO.2005.04.4875

4. Conroy T, Lamfichekh N, Etienne PL, et al. Total neoadjuvant therapy with mFOLFIRINOX versus preoperative chemoradiation in patients with locally advanced rectal cancer: Final results of PRODIGE 23 phase III trial, a UNICANCER GI trial. J Clin Oncol. 2020;38(15_suppl):4007–4007. doi:10.1200/JCO.2020.38.15_suppl.4007

5. Fernandez-Martos C, Garcia-Albeniz X, Pericay C, et al. Chemoradiation, surgery and adjuvant chemotherapy versus induction chemotherapy followed by chemoradiation and surgery: long-term results of the Spanish GCR-3 phase II randomized trial†. Ann Oncol. 2015;26(8):1722–1728. doi:10.1093/annonc/mdv223

6. Fokas E, Schlenska-Lange A, Polat B, et al. Chemoradiotherapy Plus Induction or Consolidation Chemotherapy as Total Neoadjuvant Therapy for Patients With Locally Advanced Rectal Cancer: Long-term Results of the CAO/ARO/AIO-12 Randomized Clinical Trial. JAMA Oncol. 2022;8(1):e215445. doi:10.1001/jamaoncol.2021.5445

7. Garcia-Aguilar J, Chow OS, Smith DD, et al. Effect of adding mFOLFOX6 after neoadjuvant chemoradiation in locally advanced rectal cancer: a multicentre, phase 2 trial. Lancet Oncol. 2015;16(8):957–966. doi:10.1016/S1470-2045(15)00004-2

8. Franke AJ, Parekh H, Starr JS, Tan SA, Iqbal A, George TJ. Total Neoadjuvant Therapy: A Shifting Paradigm in Locally Advanced Rectal Cancer Management. Clin Colorectal Cancer. 2018;17(1):1–12. doi:10.1016/j.clcc.2017.06.008

9. Garcia-Aguilar J, Patil S, Gollub MJ, et al. Organ Preservation in Patients With Rectal Adenocarcinoma Treated With Total Neoadjuvant Therapy. J Clin Oncol. 2022;40(23):2546–2556. doi:10.1200/JCO.22.00032

10. Conroy T, Bosset JF, Etienne PL, et al. Neoadjuvant chemotherapy with FOLFIRINOX and preoperative chemoradiotherapy for patients with locally advanced rectal cancer (UNICANCER-PRODIGE 23): a multicentre, randomised, open-label, phase 3 trial. Lancet Oncol. 2021;22(5):702–715. doi:10.1016/S1470-2045(21)00079-6

11. Dewdney A, Cunningham D. Toward the non-surgical management of locally advanced rectal cancer. Curr Oncol Rep. 2012;14(3):267–276. doi:10.1007/s11912-012-0234-z

12. Glynne-Jones R, Harrison M, Hughes R. Challenges in the neoadjuvant treatment of rectal cancer: balancing the risk of recurrence and quality of life. Cancer Radiother J Soc Francaise Radiother Oncol. 2013;17(7):675–685. doi:10.1016/j.canrad.2013.06.043

13. Maas M, Nelemans PJ, Valentini V, et al. Long-term outcome in patients with a pathological complete response after chemoradiation for rectal cancer: a pooled analysis of individual patient data. Lancet Oncol. 2010;11(9):835–844. doi:10.1016/S1470-2045(10)70172-8

14. Habr-Gama A, Perez RO, Nadalin W, et al. Operative versus nonoperative treatment for stage 0 distal rectal cancer following chemoradiation therapy: long-term results. Ann Surg. 2004;240(4):711–717; discussion 717-718. doi:10.1097/01.sla.0000141194.27992.32

15. Smith JJ, Chow OS, Gollub MJ, et al. Organ Preservation in Rectal Adenocarcinoma: a phase II randomized controlled trial evaluating 3-year disease-free survival in patients with locally advanced rectal cancer treated with chemoradiation plus induction or consolidation chemotherapy, and total mesorectal excision or nonoperative management. BMC Cancer. 2015;15(1):767. doi:10.1186/s12885-015-1632-z

16. Verheij FS, Omer DM, Williams H, et al. Long-Term Results of Organ Preservation in Patients With Rectal Adenocarcinoma Treated With Total Neoadjuvant Therapy: The Randomized Phase II OPRA Trial. J Clin Oncol. Published online October 26, 2023. doi:10.1200/JCO.23.01208

17. Wiltink LM, Nout RA, van der Voort van Zyp JRN, et al. Long-Term Health-Related Quality of Life in Patients With Rectal Cancer After Preoperative Short-Course and Long-Course (Chemo) Radiotherapy. Clin Colorectal Cancer. 2016;15(3):e93–99. doi:10.1016/j.clcc.2016.02.012

18. Hupkens BJP, Martens MH, Stoot JH, et al. Quality of Life in Rectal Cancer Patients After Chemoradiation: Watch-and-Wait Policy Versus Standard Resection - A Matched-Controlled Study. Dis Colon Rectum. 2017;60(10):1032–1040. doi:10.1097/DCR.0000000000000862

19. Quezada-Diaz FF, Smith JJ, Jimenez-Rodriguez RM, et al. Patient-Reported Bowel Function in Patients with Rectal Cancer Managed by a Watch-and-Wait Strategy after Neoadjuvant Therapy: A Case-Control Study. Dis Colon Rectum. 2020;63(7):897–902. doi:10.1097/DCR.0000000000001646

20. Smith JJ, Strombom P, Chow OS, et al. Assessment of a Watch-and-Wait Strategy for Rectal Cancer in Patients With a Complete Response After Neoadjuvant Therapy. JAMA Oncol. 2019;5(4):e185896. doi:10.1001/jamaoncol.2018.5896

21. Cremolini C, Antoniotti C, Stein A, et al. Individual Patient Data Meta-Analysis of FOLFOXIRI Plus Bevacizumab Versus Doublets Plus Bevacizumab as Initial Therapy of Unresectable Metastatic Colorectal Cancer. J Clin Oncol Off J Am Soc Clin Oncol. Published online August 20, 2020:JCO2001225. doi:10.1200/JCO.20.01225

22. Falcone A, Ricci S, Brunetti I, et al. Phase III trial of infusional fluorouracil, leucovorin, oxaliplatin, and irinotecan (FOLFOXIRI) compared with infusional fluorouracil, leucovorin, and irinotecan (FOLFIRI) as first-line treatment for metastatic colorectal cancer: the Gruppo Oncologico Nord Ovest. J Clin Oncol Off J Am Soc Clin Oncol. 2007;25(13):1670–1676. doi:10.1200/JCO.2006.09.0928

23. Loupakis F, Cremolini C, Masi G, et al. Initial therapy with FOLFOXIRI and bevacizumab for metastatic colorectal cancer. N Engl J Med. 2014;371(17):1609–1618. doi:10.1056/NEJMoa1403108

24. Cremolini C, Loupakis F, Antoniotti C, et al. FOLFOXIRI plus bevacizumab versus FOLFIRI plus bevacizumab as first-line treatment of patients with metastatic colorectal cancer: updated overall survival and molecular subgroup analyses of the open-label, phase 3 TRIBE study. Lancet Oncol. 2015;16(13):1306–1315. doi:10.1016/S1470-2045(15)00122-9

25. Gruenberger T, Bridgewater J, Chau I, et al. Bevacizumab plus mFOLFOX-6 or FOLFOXIRI in patients with initially unresectable liver metastases from colorectal cancer: the OLIVIA multinational randomised phase II trial. Ann Oncol Off J Eur Soc Med Oncol. 2015;26(4):702–708. doi:10.1093/annonc/mdu580

26. Schmoll H, Garlipp B, Junghanß C, et al. FOLFOX/bevacizumab +/- irinotecan in advanced colorectal cancer (CHARTA): Long term outcome. Ann Oncol. 2018;29:v108. doi:10.1093/annonc/mdy149.022

27. Hurwitz HI, Tan BR, Reeves JA, et al. Phase II Randomized Trial of Sequential or Concurrent FOLFOXIRI-Bevacizumab Versus FOLFOX-Bevacizumab for Metastatic Colorectal Cancer (STEAM). The Oncologist. 2019;24(7):921–932. doi:10.1634/theoncologist.2018-0344

28. Sastre J, Vieitez JM, Gomez-España MA, et al. Randomized phase III study comparing FOLFOX + bevacizumab versus folfoxiri + bevacizumab (BEV) as 1st line treatment in patients with metastatic colorectal cancer (mCRC) with ≥3 baseline circulating tumor cells (bCTCs). J Clin Oncol. 2019;37(15_suppl):3507-3507. doi:10.1200/JCO.2019.37.15_suppl.3507

29. Van Cutsem E, Cervantes A, Adam R, et al. ESMO consensus guidelines for the management of patients with metastatic colorectal cancer. Ann Oncol Off J Eur Soc Med Oncol. 2016;27(8):1386–1422. doi:10.1093/annonc/mdw235

30. Benson AB, Venook AP, Al-Hawary MM, et al. Colon Cancer, Version 2.2021, NCCN Clinical Practice Guidelines in Oncology. J Natl Compr Cancer Netw JNCCN. 2021;19(3):329–359. doi:10.6004/jnccn.2021.0012

31. Yoshino T, Arnold D, Taniguchi H, et al. Pan-Asian adapted ESMO consensus guidelines for the management of patients with metastatic colorectal cancer: a JSMO-ESMO initiative endorsed by CSCO, KACO, MOS, SSO and TOS. Ann Oncol Off J Eur Soc Med Oncol. 2018;29(1):44–70. doi:10.1093/annonc/mdx738

32. Fokas E, Allgäuer M, Polat B, et al. Randomized Phase II Trial of Chemoradiotherapy Plus Induction or Consolidation Chemotherapy as Total Neoadjuvant Therapy for Locally Advanced Rectal Cancer: CAO/ARO/AIO-12. J Clin Oncol. Published online May 31, 2019. doi:10.1200/JCO.19.00308

33. Boysen AK, Schou JV, Spindler KLG. Cell-free DNA and preoperative chemoradiotherapy for rectal cancer: a systematic review. Clin Transl Oncol Off Publ Fed Span Oncol Soc Natl Cancer Inst Mex. 2019;21(7):874–880. doi:10.1007/s12094-018-1997-y

34. Massihnia D, Pizzutilo EG, Amatu A, et al. Liquid biopsy for rectal cancer: A systematic review. Cancer Treat Rev. 2019;79:101893. doi:10.1016/j.ctrv.2019.101893

35. Morais M, Pinto DM, Machado JC, Carneiro S. ctDNA on liquid biopsy for predicting response and prognosis in locally advanced rectal cancer: A systematic review. Eur J Surg Oncol J Eur Soc Surg Oncol Br Assoc Surg Oncol. 2022;48(1):218–227. doi:10.1016/j.ejso.2021.08.034

36. Wang Y, Yang L, Bao H, et al. Utility of ctDNA in predicting response to neoadjuvant chemoradiotherapy and prognosis assessment in locally advanced rectal cancer: A prospective cohort study. PLoS Med. 2021;18(8):e1003741. doi:10.1371/journal.pmed.1003741

37. Reinert T, Henriksen TV, Christensen E, et al. Analysis of Plasma Cell-Free DNA by Ultradeep Sequencing in Patients With Stages I to III Colorectal Cancer. JAMA Oncol. 2019;5(8):1124–1131. doi:10.1001/jamaoncol.2019.0528

38. Tie J, Cohen JD, Lahouel K, et al. Circulating Tumor DNA Analysis Guiding Adjuvant Therapy in Stage II Colon Cancer. N Engl J Med. 2022;386(24):2261–2272. doi:10.1056/NEJMoa2200075

39. Tie J, Cohen JD, Wang Y, et al. Serial circulating tumour DNA analysis during multimodality treatment of locally advanced rectal cancer: a prospective biomarker study. Gut. 2019;68(4):663–671. doi:10.1136/gutjnl-2017-315852

